# Screening and managing of suspected or confirmed novel coronavirus (COVID-19) patients: experiences from a tertiary hospital outside Hubei province

**DOI:** 10.1101/2020.03.20.20038679

**Authors:** Hong Pu, Yujun Xu, Gordon S. Doig, Yan Zhou

## Abstract

**Objectives:** To report our experiences screening and managing patients with suspected or confirmed novel coronavirus (COVID-19) disease using a hospital-specific protocol.

**Design:** Longitudinal cohort study.

**Setting:** A 1,200 bed tertiary care teaching hospital in Chengdu, Sichuan, China.

**Participants:** 802 adults presenting to hospital with concerns of having COVID-19, 1,246 inpatients and 2,531 hospital visitors.

**Interventions:** Screening and management of patients using a hospital-specific protocol, which included fever triage, monitoring visitors and patients, emergency response, personnel training for healthcare team members, health education for patients and family, medical materials management, disinfection and wastes disposal protocols.

**Results:** Between 23 January and 28 February 2020, 73 people were identified as having fever plus respiratory signs with/without a history of exposure and were tested for the severe acute respiratory syndrome coronavirus 2 (SARS-CoV-2) by our hospital lab using RT PCR. Forty-five of these 73 people were subsequently excluded based on one negative RT PCR result plus positive results to quick screening tests for flu or other respiratory viruses. The remaining 28 people received a second RT PCR test 24 h later. Three people were confirmed positive for COVID-19 based on two consecutive positive RT PCR tests whilst 25 people were excluded based on two consecutive negative tests. The three COVID-19 confirmed cases received non-critical care. There were no new infections of medical staff or new infections of other hospital inpatients.

**Conclusions:** A hospital-specific protocol for screening and management is necessary for reliably identifying suspected or confirmed COVID-19 patients during an outbreak. All three cases were detected as a result of vigilant monitoring of hospital visitors. Whilst screening out-patients presenting to a fever clinic remains important, monitoring visitors must not be overlooked.

**Strengths and limitations of this study:**

► We report a hospital-specific protocol used to screen and manage people presenting to our hospital fever clinic, inpatients and visitors during an outbreak of novel coronavirus (COVID-19) pneumonia in Chengdu, Sichuan province.
► Key components of the protocol included: a three-level fever triage process; monitoring visitors and inpatients, formation of an emergency response team for COVID-19, personnel training for healthcare team members, health education for patients and family, medical materials management, and disinfection and wastes disposal protocols.
► The ability to test nucleic acid of SARS-CoV-2 using RT PCR in the hospital greatly shortened the time from the detection of patients to diagnosis, and was beneficial to the control of the transmission of the SARS-CoV-2.
► Although our process detected few patients, comparison with other processes, when they are published, will allow the identification of the optimal approach for screening and management.
► We suggest that if all resources had been focused on screening people through our fever clinic, we would have missed important in-hospital risks of transmitting COVID-19: The detection of a hospital visitor with COVID-19 led to the detection of an inpatient with COVID-19.

## INTRODUCTION

Novel coronavirus disease (COVID-19) was detected in Wuhan, Hubei Province, China in late 2019, and spread rapidly to over 110 countries and regions around the world. On January 30, 2020, the epidemic was recognized as a public health emergency of international concern by the World Health Organization, and was officially declared a pandemic on 11 March 2020. The increasing number of cases and deaths poses a major global public health challenge^1^ and has already brought a heavy burden to the economy of the world.^2^

By February 28, 2020, a total of 78,959 confirmed cases of novel coronavirus infection had been reported in China with an overall mortality rate of 3.53% (2,791 cases). ^3^ Of these, 13,052 cases were reported outside Hubei province, with an overall mortality rate of 0.85% (111 cases). The reasons for the reduced mortality outside Hubei province are unknown, however it is possible the causative virus, severe acute respiratory syndrome coronavirus 2 (SARS-CoV-2), loses pathogenicity due to passage, it is possible primary care is now more appropriate than during the early stages of the outbreak and it is also possible that screening for diagnosis and management is more appropriate.^4^

During the outbreak in Sichuan province, we developed and implemented a hospital-specific systematic process for screening, isolating and managing suspected and confirmed COVID-19 patients. The purpose of this paper is to report the effectiveness of this process.

## MATERIALS AND METHODS

### Study design and participants

We conducted a longitudinal cohort study of adults presenting to hospital with concerns of having COVID-19, inpatients and hospital visitors between 23 January and 28 February in Shang Jin Nan Fu Hospital, a tertiary care teaching hospital located in Chengdu, Sichuan province. It has 1,200 patient beds, including 50 intensive care unit (ICU) bed spaces. Five ICU bed spaces are single rooms capable of negative pressure filtration.

### Interventions

#### Three-level fever triage

A three-level fever triage system was developed. The first level of fever triage occurred at the main entrance of the emergency department and outpatient building. The second level occurred at the nurse triage station in each clinic, and the third level occurred in each clinical department, respectively. This triage system was expected to avoid missing suspected cases through repeated temperature monitoring. Temperature of every patient and his/her companion who presented to our hospital were assessed by infrared thermometer (forehead or wrist) at each level of fever triage and the epidemiological history was inquired in detail (Table 1).

**Table 1.**
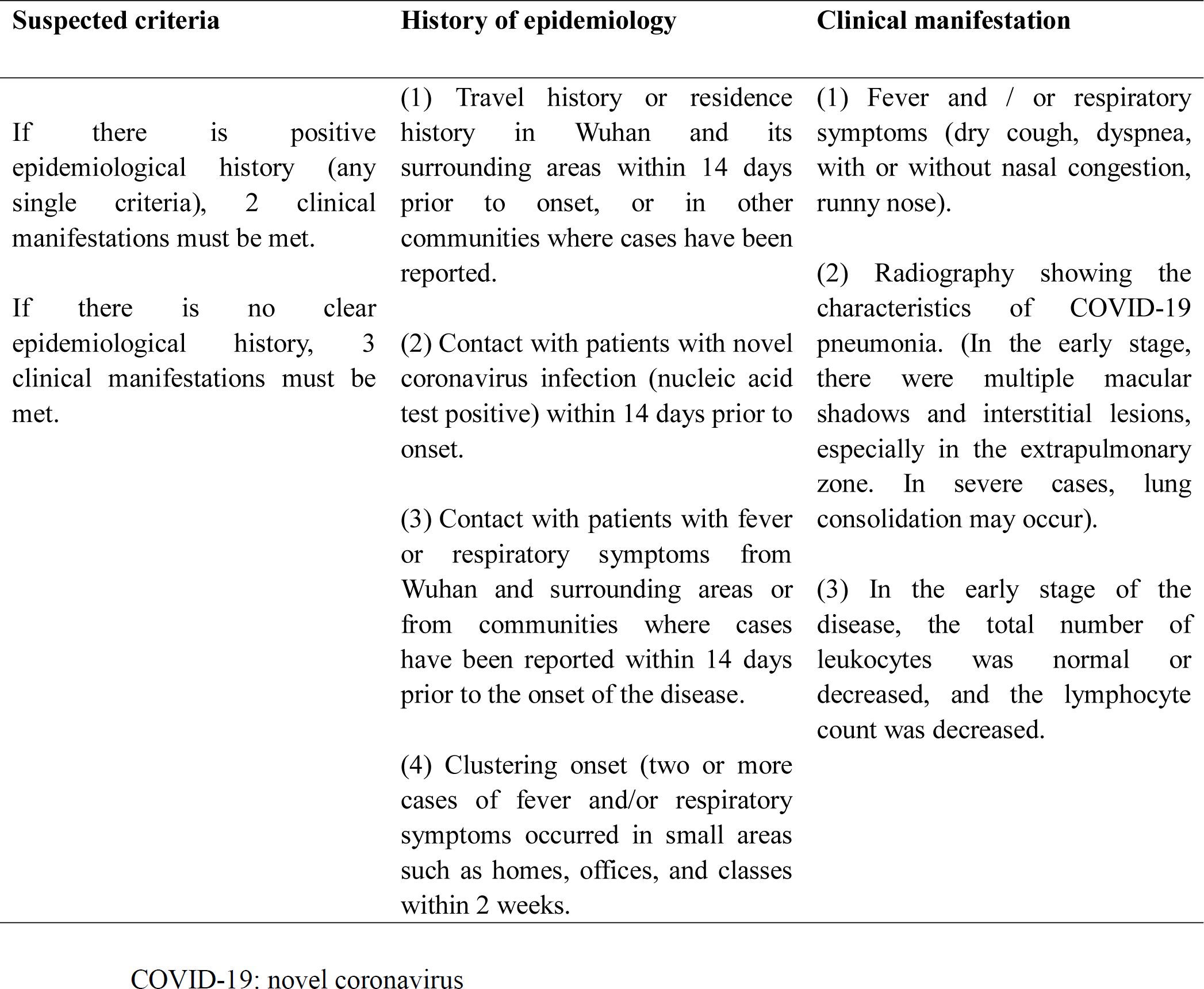
**Clinical manifestations and epidemiological history used to establish suspected case of COVID-19 pneumonia^5^**

Patients with “fever and respiratory disease” or “fever of unknown origin” were guided to visit the fever clinic which was managed by the Emergency Department. Doctors who have been trained were designated to organize the screening, consultation, referral and reporting of suspected cases.

During the outbreak, the hospital reserved a transfer channel and elevator near the prepared observational room. The channel and the elevator did not operate normally. In case of suspected or confirmed cases, the dedicated channel and elevator were used for transfer. Specially designated access for staff was opened to avoid cross infection.

#### Monitoring visitors

Visitors were strictly restricted. Each patient could only be accompanied by one person, and each accompanying person must accept daily temperature monitoring and report any symptoms of respiratory disease. Visitors were required to wear personal protective equipment and follow hand hygiene rules. Visitors of ICU patients were refused.

#### Emergency response

Once a fever patient with respiratory symptoms or a patient with fever of unknown origin plus positive history of epidemiology was identified by the fever clinic, the emergency response system was initiated immediately. Similarly, if any clinical department within our hospital identified a fever patient with respiratory symptoms or a patient with fever of unknown origin plus positive history of epidemiology was identified, the emergency response system would be initiated.

In accordance with “Diagnosis and treatment guideline for novel coronavirus pneumonia (Trial version 6)”,^3^ routine blood test, chest radiography, quick screening of swab or respiratory secretion (sputum, tracheal suction or bronchoalveolar lavage fluid) for influenza A/B virus antigen (colloidal gold method, obtained results in 20 minutes) and 13 common respiratory viruses (nucleic acid test, obtained results in 8 hours) were completed in the observational room of the fever clinic or in the isolating room of the clinical ward.

Experts in the consultation team, consisting of specialists in emergency medicine, respiratory medicine, traditional Chinese medicine, infectious disease and intensive care, were contacted to guide further examination and treatment. The radiological department provided separate portable machines and staff for suspected patients and initiated a priority channel to ensure rapid results and minimal contamination. The CT room was equipped with a continuous air disinfector (AJ/YXD-III plasma air purifier, China), making use of ultraviolet ray dynamic antivirus and ozone sterilization.

Clinically suspected patients were isolated immediately: 1) if identified by the fever clinic, the patient was isolated in the observational room of fever clinic, and then moved to a task-specific isolating room nearby; 2) if identified by a clinical department, the patient was moved to a prepared isolating room.

The swab or respiratory secretion was collected for first SARS-CoV-2 testing in our hospital lab using real-time fluorescence Reverse Transcription-Polymerase Chain Reaction (RT-PCR) in all patients having a fever plus respiratory signs with/without a history of exposure and meeting the criteria of suspected case (Table 2). The result was obtained within 6 hours in our lab.

**Table 2.** Severity classification of COVID-19 pneumonia^5^.

| **Mild** | **Common** | **Severe** | **Critical** |
| --- | --- | --- | --- |
| Low fever, mild fatigue, and there was no sign of pneumonia on radiography. | Fever and other common flu symptoms, radiography shows characteristics of pneumonia. | Any of the following:<br>(1) Dyspnea, respiratory rate $\geq 30$ times / min;<br>(2) In the resting state, the oxygen saturation is $\leq 93\%$;<br>(3) $\text{PaO}_2 / \text{FiO}_2 \leq 300\text{mmHg}$;<br>(4) Radiography shows the lesions progressed more than 50% in less than 48 hours. | Any of the following:<br>(1) Mechanical ventilation is required;<br>(2) Shock;<br>(3) Patients with other organ failure needing monitoring and treatment in an ICU. |

COVID-19: novel coronavirus

Patients were excluded from having COVID-19 based on one negative RT PCR result plus positive results to quick screening tests for flu or other respiratory viruses, negative routine blood test and radiography. Patients who could not be ruled out were sampled by the District Center for Disease Control and Prevention (District CDC), which conducted additional epidemiological investigations and collected additional samples for double SARS-CoV-2 testing 24 h later. Patients’ detailed information, diagnosis, time of collecting samples, time of returning test report, specimen number, test results and other information were recorded to provide evidence for the clinical decision.

Patients were confirmed positive for COVID-19 based on two consecutive positive RT PCR tests. Patients were excluded based on two consecutive negative test results. The excluded patients were sent to the general outpatient clinic or ward for routine treatment but were kept in isolation for at least 2 days, or discharged home for self-isolation. If the patient’s condition changed, the screening procedure was initiated again.

Confirmed non-critical patients were transferred to the local public health clinical medical service center in a dedicated vehicle with a negative pressure filtration system.

The critically ill patients were transferred to the ICU with negative pressure filtration until their condition became stable and suitable for transfer. See Table 2 for severity classification criteria of COVID-19 pneumonia.

After onset of COVID-19, a patient was determined to be ‘recovered’ when all of the following criteria were met: 1) Body temperature returned to normal for more than three days; 2) Respiratory symptoms improved significantly; 3) Chest radiography showed that the acute exudative lesions were significantly improved; 4) The RT PCR test for SARS-CoV-2 was negative for two consecutive tests obtained at least 24 hours apart. Once recovered, patients were discharged home for 14 days isolation or 14 days isolation in a special quarantine facility.

#### Personnel training

At the beginning of the outbreak, staff training on COVID-19 pneumonia was organized immediately. The main contents of the training were: knowledge of characteristics of COVID-19 pneumonia and pneumonia of unknown origin; skills in collecting history of epidemiology; diagnostic criteria, principles of treatment and requirements for reporting on outbreaks; skills of disinfection, isolation and personal protection. All medical staff took standard preventive measures.^5^ Working clothes and medical masks were worn during the medical activities and rounds. The requirement of standard protection was wearing disposable preventive clothing or with protective apron, disposable working cap, protective mask, goggles or screen, disposable latex gloves, and shoe covers if necessary.

#### Health education

The nurses in the ward carried out health education for patients and companions, including personal protection, timely reporting of respiratory symptoms, not concealing epidemiological history, minimizing contact with others, correctly implementing coughing etiquette and hand hygiene.

#### Medical material management

In order to solve the shortage of medical equipment, such as surgical masks, face shields, N95 masks, protective clothing and goggles, etc., the hospital distributed them to each department according to their needs. All medical materials were supervised by specially-assigned personnel.

#### Disinfection and wastes disposal

The medical devices, surface of the items and ground were disinfected daily with disinfectant containing chlorine 500-2000 mg/L or 75% alcohol in the fever clinic and the room with suspected or confirmed case. All wastes generated from suspected or confirmed cases were marked with “COVID-19” and treated as infectious wastes.

## RESULTS

Between 23 January and 28 February 2020, 802 people presented to the hospital’s fever clinic. In addition, 1,246 inpatients and 2,531 visitors were screened for signs of COVID-19. Seventy-three people were identified as having respiratory signs plus fever with/without a history of exposure and therefore qualified for testing for SARS-CoV-2 by our hospital lab using RT PCR.

Forty-five of these 73 patients were subsequently excluded from having COVID-19 based on one negative RT PCR result plus positive results to quick screening tests for flu or other respiratory viruses. The remaining 28 patients were sampled 24 h later for a second RT PCR test. Three patients were confirmed positive for COVID-19 based on two consecutive positive RT PCR tests whilst 25 patients were excluded based on two consecutive negative test results.

Of the three confirmed cases, one was a female visitor in her late 40’s who had no clear history of epidemiology but was identified as having cough. She was immediately sent to the fever clinic and was identified as having a fever. Through our emergency response system, she was quickly sampled for screening and confirmed COVID-19 after two consecutive positive SARS-CoV-2 tests. As a result of contact tracing, the female inpatient she visited and one male relative were subsequently confirmed positive for COVID-19 based on two positive RT PCR tests. These three COVID-19 confirmed patients were sent to the public health clinical service center for non-critical care. The male patient has been discharged after repeated negative SARS-CoV-2 testing, whilst the two female patients have not yet met discharge criteria and remained hospitalized at the end of the study follow-up period (28 February 2020).

There were no new infections of medical staff and no new infections of other inpatients in our hospital during this study period.

## DISCUSSION

Between 23 January and 28 February 2020, we screened 802 people presenting to the hospital’s fever clinic, 1,246 inpatients and 2,531 visitors through our protocol. Seven hundred and twenty-nine people identified as low risk and sent directly home. Only three patients were confirmed positive for COVID-19 based on two consecutive positive RT PCR tests, however, these patients were detected based on vigilant screening of hospital visitors.

Interestingly, active screening of 802 people presenting to our hospital’s fever clinic because they were concerned they had COVID-19 did not detect any confirmed cases during this outbreak. If we had focused all resources on screening outpatients at the fever clinic, we would have missed important in-hospital risks. We strongly recommend that screening resources should focus on self-presenting patients, hospital visitors and inpatients. Furthermore, the rapid establishment of the ability to test nucleic acid of SARS-CoV-2 using RT PCR in our hospital greatly shortened turn-around time and was beneficial in that it allowed early isolation and control of high-risk patients and discharge of low-risk (negative) patients.

A hospital-specific protocol for screening and management is necessary if an epidemic outbreak occurs. In the situation of an unanticipated surge of patients combined with a relative shortage of medical resources (beds, staff and PPE), the establishment of screening and diagnostic protocol for suspected patients can focus medical resources on patients who need them and may help reduce missed cases.

We are unaware of any other papers documenting hospital screening processes for COVID-19. Although our process detected few patients, comparison with other processes, when they are published, perhaps will allow the identification of the optimal approach for screening and management.

## Data Availability

Data are available upon reasonable request.

## Acknowledgement

The authors thank the contributions made by Yan Zhong from Hospital management committee, Shi Qiu from Department of medical administration,Shilan Xu from Department of nosocomial management, Junjun Chen, Dianrong Wang, Zhi Li, Youhui Zhao, Jianwen Wang and Fu Yang from Critical Care Department of West China Hospital and Changwei Chen from Department of Anesthesia of West China Hospital.

## Contributors

Pu conceived the study. Zhou supervised the conduct of the data collection. Xu collected and managed the data. Pu and Doig drafted the manuscript. All authors contributed substantially to its revision. Zhou takes responsibility for the paper as a whole.

## Patient public involvement

Patients and/or the public were not involved in the design, or conduct, or reporting or dissemination plans of this research.

## Patient consent for publication

Not required

## Ethics approval

Not applicable

## Provenance and peer review

Not commissioned; externally peer reviewed.

## Data availability statement

Data are available upon reasonable request.

## Open access

This is an open access article distributed in accordance with the Creative Commons Attribution Non Commercial (CC BY-NC 4.0) license, which permits others to distribute, remix, adapt, build upon this work non-commercially, and license their derivative works on different terms, provided the original work is properly cited, appropriate credit is given, any changes made indicated, and the use is non-commercial. See: http://creativecommons.org/licenses/by-nc/4.0/

